# Agriculture-urban interfaces, social vulnerability, and climate change shape West Nile virus risk across the United States

**DOI:** 10.64898/2026.07.06.26357166

**Authors:** Samantha Sambado, Váleri N. Vasquez, Mauricio Cruz-Loya, Johannah E. Farner, Rachel L. Fay, Samantha Bents, Joshua E. Lazaro, Talya Shragai, Isabel O. Delwel, Desire I. Uwera Nalukwago, Erin A. Mordecai

**Affiliations:** Department of Biology, Stanford University, Stanford, California, United States of America; Center for International Security and Cooperation, Stanford University, Stanford, California, United States of America; Division of Infectious Diseases and Geographic Medicine, Department of Medicine, Stanford University, Stanford, California, United States of America; Center for Human and Planetary Health, Stanford Woods Institute for the Environment, Stanford, California, United States of America; Department of Microbiology and Immunology, Stanford University, Stanford, California, United States of America

**Keywords:** *Culex* mosquitoes, vector-borne disease, climate warming, West Nile virus, land cover change

## Abstract

Climate and land use change are reshaping the dynamics of vector-borne diseases. West Nile virus (WNV), the most widespread zoonotic arbovirus in the United States, illustrates the need to integrate climate, land cover, and social vulnerability across heterogenous landscapes when assessing spatial risk. We present a nationwide, county-level assessment of WNV risk, using complementary statistical and mechanistic models to (1) identify socio-ecological correlates of current WNV incidence, and (2) project vector species-specific, temperature-dependent transmission suitability under mid- and late-century climate change scenarios. We find that land cover gradients, temperature-driven transmission, and both occupational and residential exposure are associated with WNV incidence, particularly in mixed urban-agricultural landscapes. Future temperature and land cover projections suggest spatially variable shifts in environmental risk, driven by divergent physiological responses among *Culex* species vectors. Our results highlight temperature and land cover as consistent, mechanistically grounded correlates of WNV risk at the national scale, while underscoring the need for refined, species-specific analyses at local levels. These insights can inform more targeted surveillance, vector control, and climate adaptation strategies. We also identify key knowledge gaps, particularly around host and vector ecology, that must be addressed to improve public health response in the face of ongoing environmental change.

## 1. Introduction

As a widespread, climate-sensitive virus transmitted across diverse ecosystems and species, West Nile virus (WNV) serves as a model system for understanding how global change reshapes zoonotic vector-borne disease risk (1,2). Its transmission involves multiple mosquito vectors and diverse avian hosts, occurring across heterogeneous environments ranging from urban centers to agricultural areas (3–5) (Figure 1). Consequently, WNV risk to humans varies spatially and temporally, even within endemic regions (6–9). Assessing spatial risk requires considering the multifaceted nature of transmission, including environmental suitability for the enzootic cycle and factors influencing spillover to humans – especially since reported cases often underestimate true infection rates due to mild infections (6,10). Integrating these components enables more effective targeting of surveillance and adaptation efforts, particularly in regions experiencing rapid environmental change (11).

**Figure 1.**
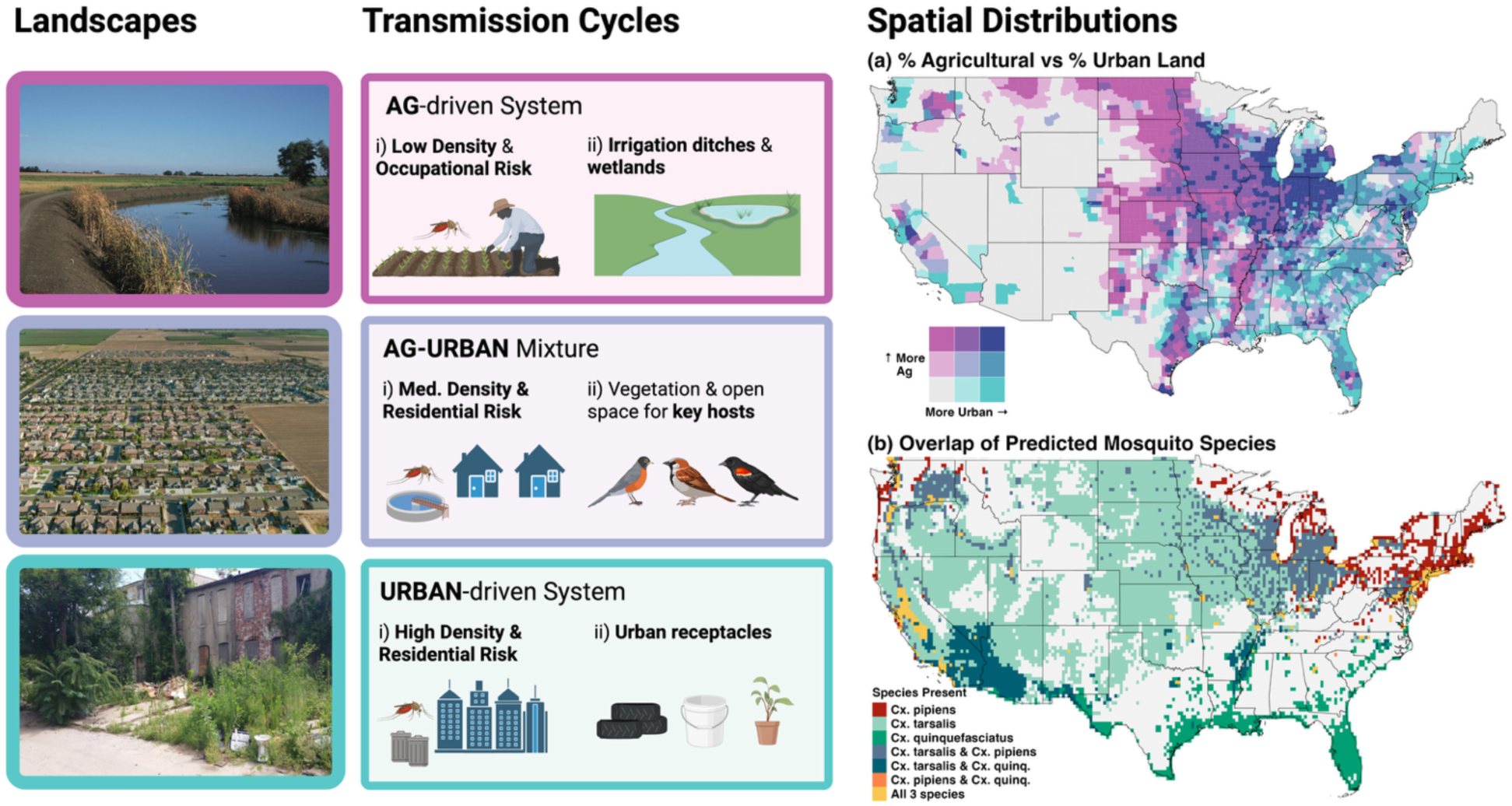
Conceptual model of WNV transmission dynamics. Transmission risk is shaped by (i) human density and (ii) mosquito-virus amplification dynamics, influenced by (a) land cover and (b) Culex species distributions. In (b), a probability threshold of ≥0.6 was used to classify pixels as occupied by a given mosquito species (32). Created with BioRender, Sambado, S (2005) https://BioRender.com/z5ck3wg. Photographs in Figure 1 (top, center, bottom) are credited to Gary Kramer (courtesy of USDA Natural Resources Conservation Science), Hal Bergman (courtesy of Getty Images), and the Baltimore Ecosystem Study LTER Photo Archive, respectively.

Previous studies have highlighted the importance of mosquito ecology (12,13), environmental drivers (2,14), and social determinants (15,16) of WNV risk, but these factors are often examined either within localized systems or through models that prioritize predictive performance using large number of potentially correlated covariates. Additionally, humans are often treated as endpoints rather than components of ecological systems, despite their role as spillover hosts whose exposure to mosquitoes varies across occupational and residential contexts. Land cover, for example, is hypothesized to influence vector-host contact rates with both humans and birds, but its effects are mediated through context-dependent ecological processes. In agricultural areas, standing water from irrigation can create ideal breeding sites for *Culex tarsalis*, potentially increasing occupational exposure for outdoor workers (6,8,17). These same irrigation ditches and waterways can also attract passerine birds, which serve as amplifying hosts for WNV (18,19). In contrast, urban environments may pose greater residential exposure to *Culex pipiens*, particularly where lower-quality housing facilitates mosquito breeding or indoor entry (20,21). Urbanization is also broadly associated with reduced bird diversity, an ecological shift that may increase human WNV risk if the birds that persist in low-diversity settings are also competent hosts (22,23). However, these coarse diversity–disease patterns may not capture complex, localized interactions between vector species and bird species of varying competence and attractiveness to vectors (13,24,25). As a result, while land cover and related variables are biologically motivated and widely used, their ability to explain spatial variation in WNV incidence at continental scales remains uncertain - particularly across heterogeneous ecological and social contexts, such as transitional zones where urban and agricultural land covers intersect and ecological interactions may become more complex (26,27).

Temperature is another key and quantifiable driver of WNV transmission, influencing mosquito and viral traits that collectively determine transmission potential (28–30). Mechanistic models of temperature-dependent transmission (i.e., R_0_(T)) capture these nonlinear relationships and can be paired with high-resolution climate and land cover projections to anticipate future environmental hotspots of disease transmission (2,29–31). In the United States (US), where multiple *Culex* species co-occur but have distinct temperature responses for transmission, species-specific modeling can clarify how warming may differentially affect vector populations (29,32). Because climate change is unlikely to affect all species uniformly, incorporating species-specific biology into risk assessment is helpful for capturing the complexity of future disease dynamics.

In this study, we address the challenge of identifying generalizable drivers of WNV risk at a continental scale by adopting a parsimonious, hypothesis-driven modeling framework. Specifically, we (1) integrate diverse socio-ecological variables to identify nonlinear correlates of current WNV incidence, and (2) project *Culex* species-specific, temperature-driven transmission suitability under mid- and late-century climate and land cover change scenarios. Our county-level results offer spatially explicit insights into how landscape structure and temperature interact to shape WNV dynamics across the contiguous US. In doing so, we highlight strategic priorities for empirical research to advance our understanding of how ecological processes can guide proactive, geographically targeted public health surveillance.

## 2. Methods

We summarize the primary modeling approaches to quantify landscape features, host-vector contact rates, and climate variability that influences WNV transmission. Full methods are provided in the Supplement (Text S1-2, Tables S1-2).

### (a) Human WNV data

County-level incidence data were obtained from ArboNET, the national surveillance system. For counties with reported cases, we used the average annual incidence of WNV neuroinvasive disease (WNVND) per 100,000 residents from 1999-2023. Counties with no case data were excluded from the analysis. Neuroinvasive cases were used due to their higher likelihood of diagnosis and reporting because cases involve more severe clinical presentations including meningitis (33).

### (b) Covariates

#### Vector habitat

We obtained county-level land cover data from the 2020 National Land Cover Database (NLCD). We defined “agricultural” land as NLCD codes 81 and 82, and “urban” land codes as 21-24. To quantify the spatial interaction between urban and agricultural areas, we calculated the urban-agricultural edge density (m/km^2^) for each county (34). Higher edge density values represent a more intermixed urban-agricultural landscape, while lower values represent more homogenous land cover. For future mosquito habitat suitability, we used projected land cover from the USGS FORE-SCE model (35) for agricultural and developed land types, and assigned species-specific suitability values for *Cx. pipiens*, *Cx. tarsalis*, and *Culex quinquefasciatus* based on known habitat associations (32).

#### Human exposure

Understanding human exposure to mosquitoes requires accounting for the diverse environmental contexts people inhabit, including occupational and residential settings. For occupational exposure, we focus on agricultural workers, who are likely at elevated risk due to prolonged outdoor activity in landscapes favorable to *Cx. tarsalis* (16). We estimated occupational exposure using county-level employment data from the Bureau of Labor Statistics Quarterly Census of Employment and Wages (QCEW). We calculated the proportion of agricultural workers relative to total private-sector employment per county. Although these data likely underestimate employment, QCEW remains the best available agricultural employment source (36). To capture potential mosquito exposure within the home environment, complementing outdoor occupational risk, we incorporated proxies that may facilitate mosquito entry, particularly for *Cx. pipiens*. Specifically, we used the CDC’s Social Vulnerability Index (SVI), which captures demographic and socioeconomic characteristics at the county level (37). Higher SVI values reflect greater vulnerability, including poverty and related social determinants that may elevate mosquito exposure risk (Text S3).

#### Bird hosts

We used the geographic ranges of 20 bird species with the highest WNV competence (based on infection studies (38)) from the USGS Gap Analysis Program to estimate total competent bird species present at the county level.

#### Climate

To estimate future climate conditions, we used temperature projections from the CMIP6 GFDL-CM4 global climate model, selected for its suitability to our study region (39). These projections were statistically downscaled to ∼6 km resolution using LOCAv2 (40). To capture multiple possible futures for the US, we analyzed two Shared Socioeconomic Pathways (SSPs): SSP2-4.5, a moderate-emission, middle-of-the-road pathway, and SSP5-8.5, a high-emission, fossil-fueled development pathway. While RCP8.5 has historically been interpreted as a “business-as-usual” scenario, recent research suggests it represents an unlikely emission trajectory (41,42). We include SSP5-8.5 to explore the upper-bound climate impacts. Climate projections were assessed for three time periods: the current period (2015–2020), mid-century (2045–2074), and late-century (2075–2100).

#### Temperature-dependent transmission

We estimated temperature-dependent transmission suitability using a mechanistic, trait-based reproductive number (R_0_(T)) model for *Cx. pipiens*, *Cx. tarsalis*, and *Cx. quinquefasciatus*, based on Mordecai et al. (2013) (43) and parameterized with WNV-specific traits from Shocket et al. (2020) (29) (Text S2, Table S2). The model incorporates temperature-dependent mosquito and viral traits, producing relative R_0_(T) values scaled 0-1 to indicate thermal suitability (hereafter referred to as R_0_(T)). To calculate annual county-level R_0_(T) values, we first calculated peak-season R_0_(T) values for each of the three *Culex* species within a focal county. To do so, for each species, we averaged monthly R_0_(T) values from the peak *Culex* activity window (April through October), accounting for potential temperature-independent diapause that occurs outside of this seasonal period. This peak-season R_0_(T) values for the three *Culex* species in a given county were then averaged at a final county-level R_0_(T) value. For temperature inputs into the R_0_(T) model, we use downscaled CMIP6 temperature projections for current, mid-, and late-century periods under two emission scenarios. We then quantified changes in thermal suitability by calculating the mean difference in R_0_(T) between current and future time periods across emission scenarios for each vector species.

### (c) Analysis

#### Current correlates of WNV

To examine how land cover, bird host dynamics, and human exposure contribute to WNV incidence, we fit a generalized additive model (GAM) using county-level average annual incidence of WNVND as the outcome (Text S2). Significant county-level covariates included urban-agricultural edge density, total competent bird species present, and proxies for human exposure (i.e., occupational and residential risk). Relative R_0_(T), based on county-level temperature data and averaged across *Culex* species, was also included as a covariate. To account for demographic differences in the risk of severe disease, we included the percentage of the population aged ≥65. All covariates were standardized prior to fitting. The outcome, WNVND incidence, was log_10_-transformed. To account for spatial autocorrelation, we included Gaussian process smooths for the latitude and longitude of county centroids. Individual covariate significance was assessed using the ‘gam.hp’ package (44). See Text S2 for model justifications and diagnostics.

#### Projected correlates of WNV

To estimate the potential for environmental WNV transmission under future climate scenarios, we developed a composite risk metric by integrating temperature-dependent transmission potential with projected mosquito habitat suitability, representing environmentally constrained transmission suitability (hereafter referred to as environmental risk). Specifically, we multiplied the species-specific R_0_(T) estimates, which are each scaled to range from 0-1, with habitat suitability layers derived from the USGS FORE-SCE land-use projections for developed and agricultural areas (35). Environmental risk values were assigned based on established habitat associations for each *Culex* species, as detailed in Text S2.3. This approach produced a scaled, spatially explicit risk metric for each species, where higher values represent areas with greater environmental risk for WNV transmission under mid- and late-century climate and land-use conditions. This approach identifies areas where both competent vector presence and thermally permissive transmission conditions overlap. Our analysis is limited to agricultural and urban land types, as these are the most ecologically relevant to the focal *Culex* species and are among the most reliably projected components of future land-use change in the US (45). We acknowledge that this simplification is a key assumption and limitation. As such, the resulting maps are intended to support visualization and exploratory spatial assessment, rather than to directly predict human disease incidence or replace localized epidemiological modeling efforts. All analyses were conducted in RStudio version 4.4.1 (46). Data cleaning and visualizations were conducted with ‘tidyversè and ‘ggplot2’ packages (47,48). Original data sources are publicly available and detailed in Table S3. Code to recreate figures and analysis are stored on Dryad repository (doi:10.5061/dryad.dz08kps9j) (49).

## 3. Results

### (a) Land cover, climate, and social vulnerability associate with current human WNV incidence

We found that county-level incidence of WNVND was significantly associated with both environmental and human exposure factors (Figure 2, Table 1). While demographic (age ≥ 65) and spatial (latitude and longitude) controls were significant covariates, our hypothesized covariates also showed consistent and ecologically plausible associations with WNV incidence. Based on the percentage of deviance explained in the full model (%), individual covariates were ranked in relative importance as follows: county longitude and latitude (75.6%), average peak season R_0_(T) (7.69%), urban-agricultural edge density (5.37%), agricultural employment (3.13%), social vulnerability index (SVI, 3.02%), percent of population aged ≥65 (2.97%), and total competent bird species (2.21%). The final model, based on 2,105 county observations, explained 72.3% of the total deviance. The large effects of spatial smoothing terms is expected given their phenomenological flexibility to capture unobserved local variation in WNV risk, which may be driven by local variation in the transmission cycle and human risk. Partial effect plots revealed non-linear relationships among the smooth terms, highlighting threshold values and offering directions for future research. The clearest threshold emerged for R_0_(T) (effective degrees of freedom, edf = 7.89) and SVI (edf = 2.92), where edf =1 indicates a linear relationship, and higher values reflect increasing non-linearity. Predicted WNV incidence trended positively with R_0_(T) but declined beyond a threshold at the upper end of the R_0_(T) range, which may reflect the upper temperature limit for *Culex* species. Risk also increased sharply when SVI exceeded ∼0.75. As a reference, counties with SVI near this threshold that are also densely populated include Queens, NY; Milwaukee, WI; and DeKalb, GA (Text S2). These findings suggest that socially vulnerable communities may face disproportionately higher WNV incidence. However, this index encompasses multiple dimensions of vulnerability (e.g., poverty, housing), so the underlying mechanisms driving this threshold remain unclear (Text S3). The relationship for urban-agricultural edge density (edf = 6.08) was also strongly non-linear, but less intuitive. Risk for WNV was highest in low edge densities, indicating that homogenous land cover – whether predominantly urban, agricultural, or another type – may promote transmission. There was a noticeable additional peak at intermediate values, suggesting increased risk in transitional zones, where urban and agricultural land cover mix. Other variables had clear linear relationships with WNV incidence: agricultural employment was positively associated, while the total number of competent bird species showed a negative relationship, potentially reflecting a disconnect between bird species richness and species evenness, which may contribute more directly to the transmission cycle. These underscore the need for mechanistic studies that also account for temporal dynamics. Together, these results suggest that current WNV transmission risk is influenced by nonlinear and interacting relationships between socio-ecological factors.

**Figure 2.**
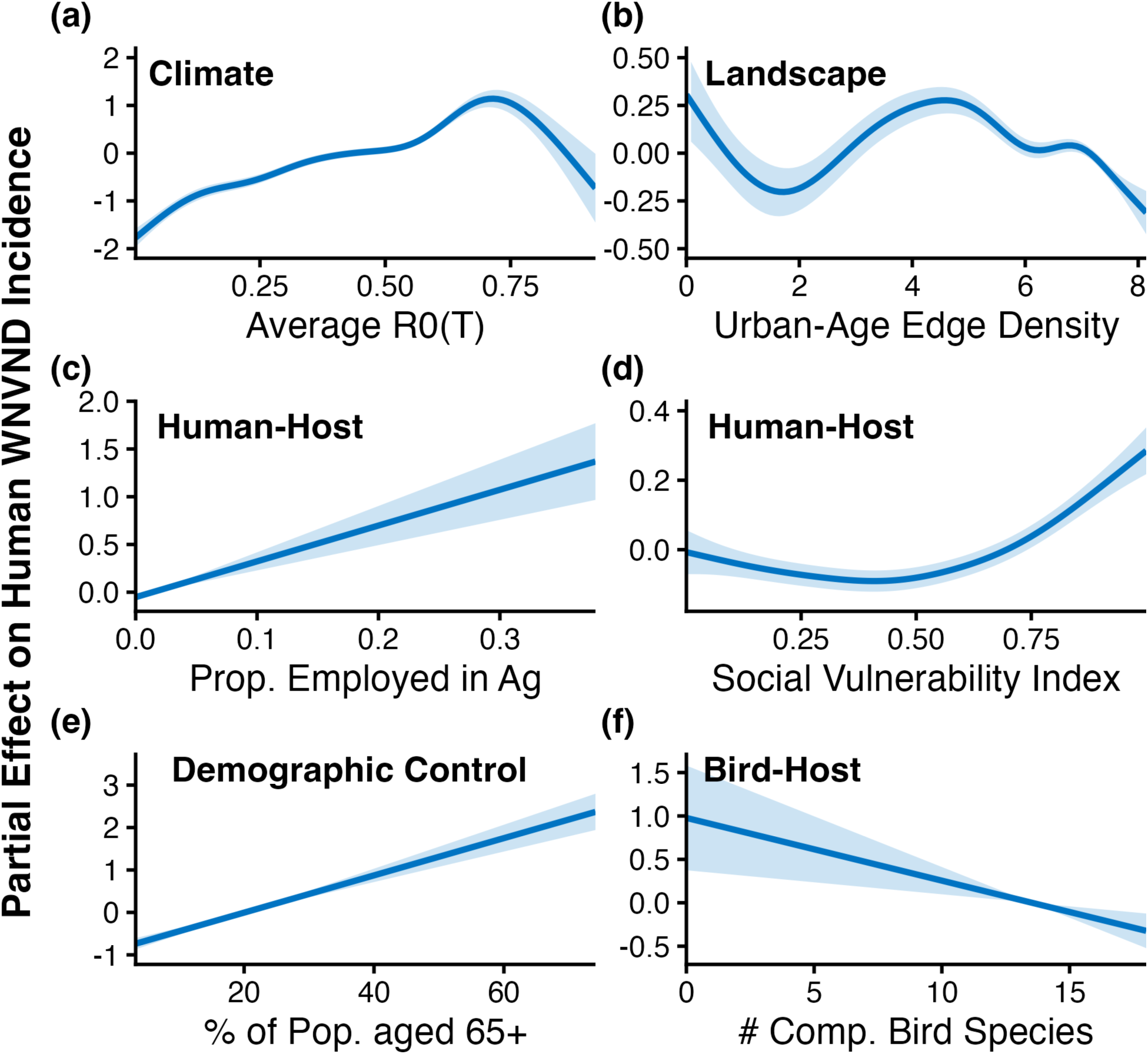
Socio-ecological correlates of current WNV incidence. Partial effects from a GAM, ordered by importance (a-f), illustrate associations between county-level WNVND incidence and covariates. Shaded areas represent 95% confidence intervals.

**Table 1.** GAM results for county-level WNV incidence. The percentage of deviance explained (%) indicates the proportion of the model’s deviance attributed to each covariate, as estimated using the ‘gam.hp’ package. Note that all covariates were initially modeled as splines, but any covariates with EDF < 2 (indicating a relationship that is approximately linear) were then fit as linear terms. Linear terms are reported as estimated coefficients with standard errors (SE), t-values, and p-values. Smooth terms are summarized by estimated degrees of freedom (edf), reference degrees of freedom (df), F-statistic, and p-values.

| Covariate | Estimate | SE | t-value | p-value | % |
| --- | --- | --- | --- | --- | --- |
| Intercept | -0.79 | 0.023 | -34.23 | $< 2.00 \times 10^{-16}$ | |
| Prop. employed in ag | 0.12 | 0.019 | 6.68 | $3.17 \times 10^{-11}$ | 3.34 |
| % of population aged $\geq 65$ | 0.22 | 0.020 | 10.86 | $< 2.00 \times 10^{-16}$ | 2.97 |
| Total competent bird species | -0.22 | 0.068 | -3.18 | 0.002 | 2.14 |
|  | EDF | DF | F-statistic | p-value | % |
| s(longitude, latitude)<sup>b</sup> | 27.58 | 19.17 | 109.43 | $< 2.00 \times 10^{-16}$ | 75.61 |
| s(average peak-season $R_0(T)$)<sup>a</sup> | 7.89 | 8.68 | 20.00 | $< 2.00 \times 10^{-16}$ | 7.94 |
| s(urban-agricultural edge density)<sup>a</sup> | 6.09 | 7.27 | 4.64 | $3.22 \times 10^{-5}$ | 5.58 |
| s(social vulnerability index)<sup>a</sup> | 2.92 | 3.65 | 6.82 | $5.67 \times 10^{-5}$ | 3.02 |

Model fit: $n = 2,105$ ; adjusted $R^2 = 0.72$ ; deviance explained = 72.3%
<sup>a</sup> cubic spline smooth; <sup>b</sup>Gaussian process smooth

### (b) Projected temperature-driven and focal land cover changes in WNV transmission

Temperature-driven transmission suitability, measured by R_0_(T), and urban-agricultural edge density emerged as the strongest mechanistic covariates in the GAM (i.e., excluding the spatial smoothing term) and are both projected to increase under most climate and land-use scenarios. However, the magnitude of change varies by mosquito species and geographic region. Across all US counties and *Culex* species, average R_0_(T) values are projected to either increase or remain stable from current to future periods under both emission scenarios (Table 2). *Cx. pipiens* shows the largest overall gain in average thermal suitability, with its mean relative R_0_(T) increasing by 0.21 (on a scale from 0 to 1). In contrast, *Cx. quinquefasciatus* and *Cx. tarsalis* exhibit more modest gains of 0.15 and 0.06, respectively. Notably, under the high-emission scenario, both *Cx. quinquefasciatus* and *Cx. tarsalis* show declines in mean R_0_(T) from mid- to late-century, indicating potential thermal constraint at elevated temperatures. By contrast, *Cx. pipiens* continues to gain thermal suitability across both future periods, reaching its peak average R_0_(T) under late-century, high-emission conditions. Spatial projections of environmental risk (Figure 3) for urban and agricultural areas reveal more nuanced, species-specific responses, which we expand upon in the Discussion. These responses underscore the importance of integrating both climate and land-use projections when anticipating future WNV risk, as environmental suitability for vectors will not shift uniformly across landscapes.

**Figure 3.**
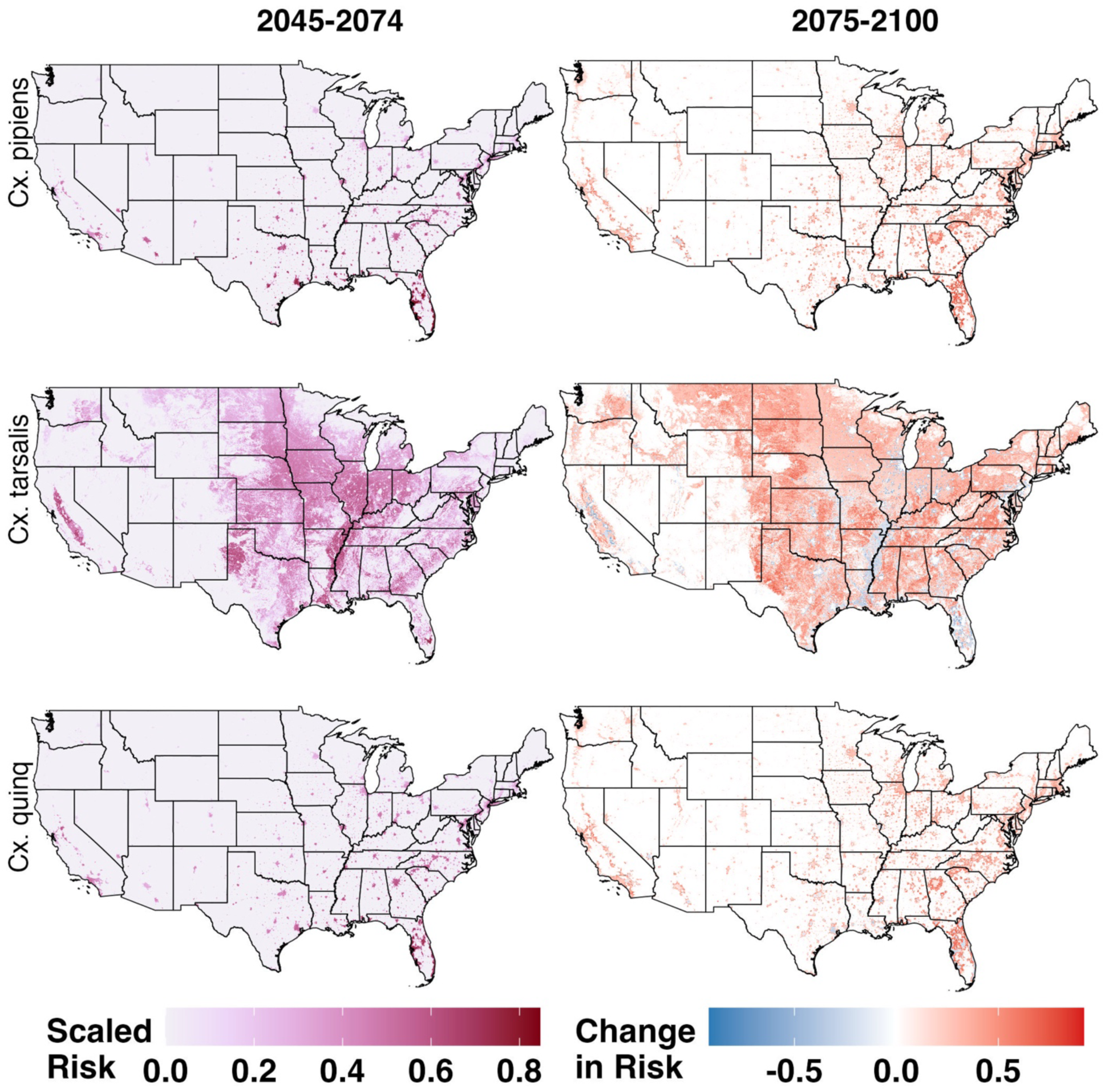
Spatial patterns of projected WNV risk. Projected scaled risk under future temperature and land-use types, based on forecasted land covers from the USGS FORE-SCE model (35) and future R0(T) estimates under the SSP2-4.5 scenario for mid-century (2045–2075) and late-century (2075–2100). Projections focus only on urban and agricultural land cover classes. Change in risk are shown relative to 2045-2074 baseline period. Rows represent *Cx. pipiens*, *Cx. tarsalis*, and *Cx. quinquefasciatus* in the top, middle, and bottom rows, respectively.

**Table 2.** Average temperature-driven transmission suitability, R_0_(T) values, across all continental US counties during peak transmission season (April through October) for each *Culex* species under current and future climate scenarios. Projections are for mid-century (2045-2074) and late-century (2075-2100) under two emission scenarios (SSP2-4.5 and SSP5-8.5). Values (mean ± standard deviation) reflect species specific thermal suitability across all counties, based on CMIP6 climate model estimates.

| Species | Current | SSP2-4.5<br>(intermediate-emission) |  | SSP5-8.5<br>(high-emission) |  |
| --- | --- | --- | --- | --- | --- |
|  |  | Mid | Late | Mid | Late |
| *Cx. pipiens* | 0.36 ± 0.22 | 0.45 ± 0.23 | 0.49 ± 0.22 | 0.50 ± 0.22 | 0.57 ± 0.19 |
| *Cx. tarsalis* | 0.48 ± 0.22 | 0.53 ± 0.20 | 0.48 ± 0.18 | 0.54 ± 0.17 | 0.51 ± 0.15 |
| *Cx. quinquefasciatus* | 0.40 ± 0.22 | 0.46 ± 0.21 | 0.55 ± 0.18 | 0.48 ± 0.18 | 0.47 ± 0.15 |

## 4. Discussion

As climate and land-use change accelerate, understanding the ecological and social factors that shape WNV transmission is increasingly critical for anticipating and mitigating emerging public health threats. Our study demonstrates that temperature and landscape structure, particularly homogenous or transitional zones between urban and agricultural areas, have a consistent but complex relationship with WNV risk. We suggest that these mixed-use environments support overlapping distributions of vectors, birds, and humans, especially in vulnerable communities (4,26,27,50). By investigating current socio-ecological indicators alongside projected species-specific, land use and temperature-dependent transmission model, we present a biologically grounded and complementary framework for understanding disease risk in a changing environment.

Our models show that land-use heterogeneity, suitable temperature for transmission, and human exposure collectively contribute to spatial variation in WNV incidence across US counties. While earlier studies have identified elevated risk in semi-urban or peri-urban zones, our work is among the first to examine national-scale edge effects, supporting prior regional results that WNV incidence is elevated in mixed environments (20,26,27,50). We hypothesize that these mixed zones combine suitable habitat, moderately competent bird hosts, and high enough human population density to drive focal WNV hotspots (24,26). We also find high risk in more homogenous landscapes, likely mirroring the importance of occupational and socioeconomic factors (15–17). Agricultural employment, a proxy for outdoor occupational exposure to *Cx. tarsalis*, showed a positive association with WNV, though further research is needed to understand underreporting and access to care in these populations (16). In urban settings, our findings align with established associations between socioeconomic disadvantage and WNV incidence (15,16,21), potentially mediated by local conditions such as poor drainage, waste accumulation, inadequate housing structure, and limited vector control, all conducive to *Cx. pipiens* proliferation (21,51). While our social vulnerability index detected a risk threshold, its aggregated nature limits mechanistic interpretation (Text S4). Subcounty analyses incorporating specific housing metrics (e.g., infrastructure age, air conditioning access) would likely improve explanatory power, especially in counties with high inequality (15,52). Prior insights from economically diverse Orange County, California underscores the importance of incorporating fine-scale socioeconomic and infrastructure into WNV incidence models, as disparities in infrastructure maintenance, such as the prevalence of neglected swimming pools, can significantly amplify disease risk in lower-income communities (15). Our bird-related findings align with previous research, suggesting that greater richness of competent bird species may be associated with reduced human WNV incidence (22,53). However, our bird metric is limited in that it does not account for seasonal abundance or host-feeding preferences of *Culex* mosquitoes under natural conditions. For example, while both American robins and American crows are competent hosts in laboratory settings, robins are more frequently fed upon by *Culex* mosquitoes in the field (25). We highlight the host-feeding preference of *Culex* species as important areas for future research to aid in our understanding of WNV dynamics (Table 3).

**Table 3.**
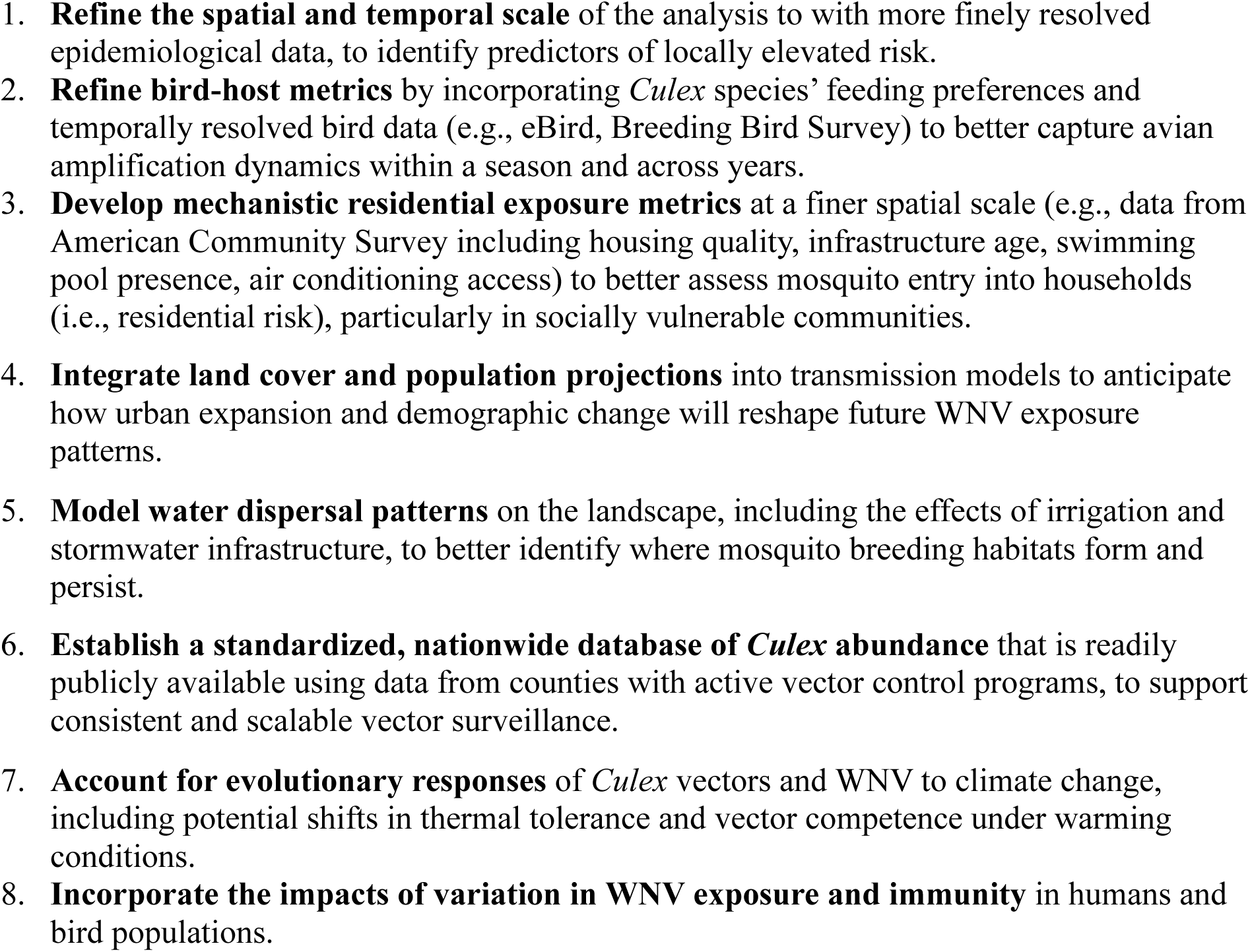
Future research priorities to improve spatial models of WNV risk under climate and land-use change.

Future WNV risk is unlikely to be spatially uniform, as each *Culex* species, and important bird hosts, respond differently to climate and land-use change due to distinct thermal physiologies and habitat preferences (54). While national averages of temperature-dependent transmission potential suggest broad trends over time (Table 2), these summaries can obscure important regional heterogeneity. Spatial projections of environmental risk (Figure 3) across urban and agricultural areas reveal nuanced, species-specific patterns. Much of the nation’s agricultural land is projected to remain thermally suitable; however, *Cx. tarsalis*, which as the broadest thermal tolerance (20.1°C), may experience localized declines in habitat suitability, particularly in regions like the Central Plains where agricultural land is projected to be converted. *Cx. quinquefasciatus*, common in the southern US, has the narrowest thermal breadth (12.7°C) and is expected to see reduced thermal suitability in parts of the south, including current endemic hotspots such as California and Arizona, where future temperatures may exceed its upper thermal limit (31.8°C) (12). *Cx. pipiens*, which has the highest upper thermal limit (34.9°C) and is widely distributed in northern urban areas, is projected to expand its range, particularly in areas undergoing urban development (30,31,55,56). These species-specific responses emphasize the importance of incorporating both climate and land-use projections when assessing future WNV risk, as environmental risk depends on the interaction between temperature thresholds and habitat availability. While our projections assume fixed thermal limits, evolutionary adaptation or behavioral plasticity in mosquitoes could shift these outcomes over time (57). The resulting county-level risk maps, which integrate temperature-dependent transmission and vector habitat suitability, provide a spatially explicit tool to support targeted surveillance and vector control under future environmental change.

From a public health perspective, our findings highlight further research directions to identify strong mechanistic links, particularly involving social vulnerability and occupational exposure, that can inform local interventions. Our results suggest that improving infrastructure in residential areas and addressing occupational risk could meaningfully reduce environmental WNV exposure, even as climate suitability increases in the US (58). Practical interventions such as improving water management, upgrading housing infrastructure, and conducting outreach to at-risk occupational groups, especially agricultural workers, may help mitigate spillover in ecologically suitable regions (15,52). Targeted management of agricultural landscapes also offer opportunities to reduce risk, for example, by optimizing irrigation practices to minimize standing water and reduce mosquito breeding habitat, particularly in arid regions where irrigation can sustain vector populations well beyond the rainy season (7,8). As urbanization continues to reshape the American landscape, there is a real risk that WNV transmission could be exacerbated without intentional, proactive planning (23,45). Newly urbanized areas, particularly in the Central Great Plains, are of special concern for *Cx. pipiens*-driven transmission given their overlap with existing WNV hotspots (4,10,17). As projected declines in agricultural land cover may reduce habitat suitability for *Cx. tarsalis*, concurrent urbanization and warming temperatures are likely to favor *Cx. pipiens*, indicating a shift in dominant transmission pathways rather than a uniform change in overall environmental suitability. In these regions, urban design that limits mosquito breeding habitat could reduce disease risk (26,51,58). Our results show the value of integrating socio-ecological and climatic data to develop forward-looking, locally-grounded public health strategies in the face of ongoing environmental change.

While our study offers novel insights into the spatial ecology of WNV on a county-level scale across the US, several caveats and limitations must be acknowledged. First, our analyses parse risk of WNVND incidence at the county level on average across the period since the virus has invaded the US—a scale too coarse to capture local relationships between land cover, birds, vectors, and humans and to capture weather anomalies that drive temporal variation. This likely explains the dominance of the spatial term and weaker model performance in regions such as the Rocky Mountains and Central Plains, where temporally dynamic processes (e.g., water availability and bird migration) play a key role. Although drought is an important climatic driver of WNV risk, it was not significant in our final analysis. This is likely because we included spatial controls that could encompass local drought variability. Drought could play a more important role in models that explicitly incorporate its temporal variability and use finer (subcounty) spatial resolution to represent vector-host dynamics (2). Notably, our static bird metric does not capture seasonal dynamics of bird migration or mosquito feeding patterns, which is important for focal WNV hotspots (24,25). Mosquito feeding behavior is another critical consideration (Text S1); however, empirical data are limited and highly localized. In addition, our use of the social vulnerability index, while informative for hypothesis generation, cannot identify mechanisms of exposure. We examined more targeted indicators that directly relate to physical conditions (Text S3) but found the county scale to be too coarse to capture the localized influence of these variables. Finally, our future projections assume a limited number of climate scenarios, and no changes in bird or human exposure metrics, which introduce uncertainty; therefore, our results should be interpretative as relative, not absolute, risk estimates.

Our findings highlight the value of mechanistic, landscape-informed models that integrate species ecology and socio-economic context to understand WNV incidence. Because the majority of WNV infections are subclinical, scalable socio-ecological frameworks are essential for proactive prediction and intervention. Advances in high-resolution climate and land-use projections, when paired with local metrics of human exposure, can refine vector control strategies under future change. Continued investment in interdisciplinary approaches will be essential to reducing the burden of WNV and other vector-borne diseases.

## Data Availability

All data are publicly available and information regarding these data can be found on a public data repository (https://doi.org/10.5061/dryad.dz08kps9j).

