## Supplementary Information for "Agriculture-urban interfaces, social vulnerability, and climate change shape West Nile virus risk across the United States"

**Table of Contents**

**Supplementary Text 1**. Data justifications and extractions
**Supplementary Text 2**. Analysis justifications
**Supplementary Text 3.** Estimating social vulnerability risk
**Supplementary Table 1**. Regional classifications
**Supplementary Table 2**. $R_0(T)$ parameters
**Supplementary Table 3.** Additional information on data availability
**Supplementary References**

SUPPLEMENTARY TEXT

**Supplementary Text 1**. Data justifications and extractions

*S Text 1.1 Data for Generalized additive models (GAMs)*

**County and regional boundaries**. County-level boundary data for the contiguous US were obtained from the 2020 US Census Bureau's TIGER/Line shapefiles using the `tigris` package (1). These shapefiles defined the geographic boundaries used for all spatial extraction and analyses conducted in this study. To facilitate regional comparisons in earlier versions of our GAMs, counties were grouped into four regions – East-Central, Southeastern, Southwestern, or Western – based on the vector geographic classification scheme used by Rochlin et al. (2019)(2), which was originally derived from vector data presented by Darsie and Ward (2005)(3). This region-specific framework supports ecological consistency with prior vector surveillance efforts (4). For our spatial autocorrelation control, we use the latitude and longitude of each county centroid which we obtained with the st_centroid() function from the `sf` package (5).

**WNV incidence**. County-level human West Nile virus (WNV) incidence data were obtained from ArboNET, the national passive surveillance system maintained by the US Centers for Disease Control and Prevention (CDC) (https://www.cdc.gov/west-nile-virus/data-maps/historic-data.html). For each county, we used the average annual incidence of reported neuroinvasive WNV disease (WNVND) per 100,000 residents from 1999-2023, based on county of residence. We focus on neuroinvasive cases because they are more likely to be diagnosed and reported (symptom manifestations include meningitis, encephalitis, or acute flaccid paralysis) compared to non-neuroinvasive WNV infections, which are often underreported due to mild or asymptomatic presentation (~70-80% of WNV infections are asymptomatic) (6). As a result, neuroinvasive incidence serves as a more reliable indicator of WNV activity for the spatial scale of our study, despite only representing a small fraction of total WNV cases. Prior studies have demonstrated a strong correlation between total WNV case counts and neuroinvasive case trends, supporting the use of this metric as a proxy for broader WNV transmission patterns (7). Nevertheless, all reported incidence is likely an underestimate, given the dependence on clinical recognition, laboratory confirmation, and reporting to public health authorities (6,8). To account for demographic factors that may increase vulnerability to neuroinvasive WNV infection, we use the percentage of population aged 65 and older at the county level. County-level estimates were obtained from the U.S. Census Bureau's American Community Survey (ACS) (Table ID: S0101, https://data.census.gov/table/ACSST5Y2020.S0101?q=S0101&g=010XX00US$0500000). These data are based on 2015-2020 ACS 5-year estimates.

**Current land cover.** County-level land cover data were obtained from the 2020 National Land Cover Database (NLCD), which provides 30 m resolution raster data with a 16-class land cover classification (https://www.mrlc.gov/data/land-cover-conus-6). For this study, land was classified as "urban" if it had an NLCD code of 21, 22, 23, or 24 (developed open space, developed low intensity, developed medium intensity, developed high intensity), and "agricultural" if it had a code of 81 or 82 (hay/pasture, cultivated crops). To calculate the percentage of each county's land that is urban or agricultural, we used the exact_extract() function from the `exactexactr` package (9). This method accounts for the fraction of each raster cell that intersects a county boundary. For each county, we summed the coverage fraction of cells that matched the relevant NLCD codes and divided by the total coverage fraction of all land cover cells in the county. This approach yields an area-weighted estimate of the proportion of land classified as urban or agricultural within each county. To create a bivariate map highlighting the relative presence of both urban and agricultural land cover, we classified each land cover type into three equal-interval bins representing low, medium, and high percentages within each county. This binning allowed us to construct a 3x3 bivariate color matrix, capturing combinations of urban and agricultural intensity across counties (Fig. 1a). To quantify the spatial interaction between urban and agricultural areas, we calculated the urban-agricultural edge density ($m/km^2$) for each county. To do this, we first created two binary rasters representing urban and agricultural land classes. Using a 3x3 moving window focal sum operation on the NLCD raster, we identified urban pixels that were adjacent to agricultural pixels. Specifically, an urban pixel was classified as an edge pixel if at least one of its eight neighboring pixels was agricultural. We then calculated the number of edge pixels within each county and multiplied by 30 m to represent NLCD pixel resolution and get total edge length per county. Edge density was standardized by dividing the total edge length by county area. Higher edge density values represent a more intermixed urban-agricultural landscape, where lower values suggest more homogenous land cover (could be urban or agricultural). This method approximates edge length using pixel-based adjacency and may overestimate true edge length in highly fragmented landscapes, particularly where corner adjacency occurs. Our goal was to capture the relative differences in landscape intermixing among counties rather than exact geometric edge length. Future studies could improve upon this method, especially in fine-scale investigations.

**Bird community competence**. Geographic layers representing species ranges and predicted habitats were obtained from the US Geological Survey (USGS) Gap Analysis Program (GAP) (https://gapanalysis.usgs.gov/apps/species-data-download/). We downloaded data for the top 20 bird species with the highest host-competence indices for West Nile virus, based on experimental infection studies (10). For each species, the GAP spatial files were reprojected to match the CRS of the county boundary shapefile. We then assessed whether each species' range intersected with individual county boundaries. If an intersection was found, the county was assigned a value of 1 for that species; otherwise it was assigned a 0 (11). This process was repeated for all 20 bird species, and the values were summed to calculate the total number of competent bird hosts present in each county (i.e., competent bird richness). However, not all competent bird species are equally fed upon by *Culex* mosquito species (12,13). As such, this metric is static and should be improved upon by including more dynamic dimensions such as varying bird abundances and seasonality of *Culex* feeding patterns (12).

**Occupational risk**. County-level employment data were obtained from the Bureau of Labor Statistics (BLS) Quarterly Census of Employment and Wages (QCEW), which provides monthly employment data categorized by industry according to the North American Industry Classification System (NAICS) (https://data.bls.gov/cew/apps/data_views/data_views.htm#tab=Tables). For this study, we focus on two specific agricultural industries identified by their NAICS code: 111 (crop production) and 1151 (support activities for crop production). The analysis was conducted using data for the years 2020-2023, the most recently available data online, and specifically for the private sector (as defined by ownership code 5). For each county, we combined the annual employment data from both NAICS 111 and NAICS 1151 to obtain the total agricultural employment per year. To provide a basis for comparison, we used the total private-sector employment across all industries (NAICS 10) per year. Next, we calculated the proportion of the workforce employed in agriculture per county by dividing the total agricultural employment (sum of NAICS 111 and

NAICS 1151) by the total private-sector employment (NAICS 10). To ensure data quality, we handled missing values by setting them to 0 for counties with no reported agricultural employment. The BLS estimates that the QCEW data represents only half of all agricultural workers since it excludes unincorporated self-employed, unpaid family members, and farm workers not covered by State insurance law, including undocumented workers (14). As such, our study underestimates the absolute number of workers with occupational risk but the QCEW remains the best available county-level employment source (15).

**Residential risk**. County-level social vulnerability data were obtained from the Centers for Disease Control and Prevention (CDC) (https://www.atsdr.cdc.gov/place-health/php/svi/index.html). Specifically, we used the Social Vulnerability Index (SVI), which captures demographic and socioeconomic characteristics at the county level. Higher SVI values reflect greater vulnerability, including poverty and related social determinants that may elevate mosquito exposure risk. The current CDC SVI uses 16 US Census variables from the 5-year American Community Survey (ACS) to identify communities that may need additional support during natural disasters. We take the average SVI from the years 2018, 2020, and 2022.

S Text 1.2 Data for projected scaled metric

**Projected climate variables**. To estimate future climate variables, we used temperature projections from the Coupled Model Intercomparison Project 6 (CMIP6), specifically the GFDL-CM4 global climate model (16), statistically downscaled to ~6 km resolution using LOCA version 2 (17). We analyzed two Shared Socioeconomic Pathway (SSP) scenarios: SSP2-4.5 (a moderate, middle-of-the-road pathway) and SSP5-8.5 (a high, fossil-fueled development pathway), and three time periods: current (2015-2020), mid-century (2045-2074), and late-century (2075-2100). For each county polygon, mean monthly raster values of minimum and maximum temperatures were extracted by time period and SSP scenario, followed by calculation of mean monthly and annual temperatures for each combination.

**Projected land use variables**. To assess how changes in land cover may influence mosquito distribution, we used projected land-cover data developed by the USGS Earth Resources Observation and Science (EROS) Center (Supplementary Text 3). These land-cover projections were modeled for four scenarios consistent with the IPCC Special Report on Emission Scenarios and downscaled using the forecasting scenarios of land-use change (FORE-SCE) model (18). These scenarios incorporate a range of socioeconomic factors – including demographics, energy consumption, and agricultural economics – to simulate anthropogenic land-use change. For this study, we selected the A2 scenario, which reflects a future characterized by regionally oriented economic development and uneven growth, closely resembling current global trends. The land-cover data were classified into 17 land-use land-cover categories, comparable to those used in the NLCD, and provided at a spatial resolution of 250 m. Projected land cover data for the years 2050 and 2100 were used to estimate habitat suitability for three mosquito species: *Cx. tarsalis*, *Cx. pipiens*, and *Cx. quinquefasciatus*. Each land-use land-cover category was assigned a habitat preference value of 0 or 0.9, with higher values indicating a greater likelihood of suitability for a given species. These values were informed by an extensive review of the literature on mosquito habitat preferences and land cover associations (Gorris et al. (2021) Table 2) (19). For example, *Cx. tarsalis* is more commonly associated with cultivated crop areas than with land cover used as mining sites or perennial ice/snow (assigned 0.9 suitability for agricultural land types (codes 12, 14). On the other hand, *Cx. pipiens* and *Cx. quinquefasciatus* is more commonly associated with developed land rather than evergreen forest (assigned 0.9 suitability for developed land types (code 2). Using these species-specific preference values, categorical land-use land-cover categories were reclassified into continuous habitat suitability raster for each mosquito species. For mapping purposes, we reprojected the raster layer to ~1.5 km resolution.

To simplify Figure 3, we focus only on projected land cover changes for urban (i.e., developed) and agricultural areas, the two categories with the highest certainty and expected rates under urbanization scenarios in the continental US. This decision also better aligns with our edge density metric, as both urban and agricultural land uses are known to influence WNV transmission. In Figure 3, the projected panel shows the scaled risk (0-1) for projected $R_0(T)$ and land cover (only urban and agricultural land cover types) in 2050. The 2100 (2075-2100) maps show the absolute change in scaled risk relative to 2050 (2045-2074), offering a clearer visualization of late-century trends. For projected environmental risk we use USGS FORE-SCE model (18) and based the relative change in risk from 2075-100 to 20245-2074.

**Supplementary Text 2**. Analysis justifications

*S Text 2.1 Generalized additive models (GAMs)*

Many efforts were made to develop and validate the final GAM presented in the main text. To determine the appropriate functional form of each covariate, we initially fit a full model with all terms specified as smooth functions. We then examined the estimated degrees of freedom (edf) for each term; covariates with edf > 2 were retained as smooth terms, while others were modeled linearly. To check if the number of years influenced our GAM results, we ran an additional model where we included the number of years of reported cases between 1999-2023. To calculate the number of years of reported cases per county, we downloaded individual years of WNV neuroinvasive case data per county from ArboNET, and identified the first year of a reported case. Results from this model were very similar to our GAM using the more standardized county incidence data from ArboNET. We also explored multiple approaches to spatial control, including regional identities (e.g., East-Central, Southwestern) and geographic coordinates (latitude and longitude), as detailed below.

Additional analyses – spatial smoothers: The table below presents the results from GAMs using different spatial control strategies, while keeping the hypothesized covariates (Term) consistent across models. Specifically, we compared three models (Model): (1) no spatial term, (2) a categorical regional spatial term based on Rochlin et al. (2019), and (3) continuous spatial control using county centroid latitude and longitude (the final model used in the main manuscript). For each model, we report the total deviance explained by the full model (under Model column), as well as the relative percentage of  deviance explained by individual predictors within the model (% explained by individual term). Individual term contributions were calculated using the `gam.hp` package (20).

| Model | Term | % explained by individual term |
|---|---|---|
| **No spatial component** | **$R_0(T)$** | 33.36 |
| | **Edge Density** | 24.19 |
| % deviance explained by full model = **29.5%** | SVI | 12.01 |
| | Ag Employment | 11.98 |
| | Total Comp. Birds | 9.64 |
| | Pop. Aged ≥ 65 | 8.82 |
| **Regional spatial component** | **Region** | 50.02 |
| | **$R_0(T)$** | 21.12 |
| % deviance explained by full model = **47.9%** | **Edge Density** | 8.82 |
| | Pop. Aged ≥ 65 | 5.22 |
| | SVI | 5.06 |
| | Ag Employment | 4.91 |
| | Total Comp. Birds | 4.85 |
| **Lat, Lon spatial component** | **Lat, lon** | 75.61 |
| | **$R_0(T)$** | 7.94 |
| % deviance explained by full model = **72.3%** | **Edge Density** | 5.58 |
| | Ag Employment | 3.34 |
| | SVI | 3.02 |
| | Pop. Aged ≥ 65 | 2.97 |
| | Total Comp. Birds | 2.14 |

Additional analyses – regional effects: To identify region-specific patterns in the relationship between predictors and human WNV neuroinvasive disease, we subset the dataset by regional identity – East Central, Southeastern, Southwestern, Western – and fit separate GAMs for each region (Model). Spatial controls were not included in these models. The table below presents the results of these region-specific models, with *N* representing the number of counties within each region. For each model, we report the total deviance explained by the full model, along with the percentage of deviance explained by each individual predictors (Term) within the model (% explained by individual term). Individual term contributions were calculated using the `gam.hp` package (20).

| **Model** | **Term** | **% explained by individual term** |
|---|---|---|
| **East Central** | **Edge Density** | 34.88 |
| | **$R_0$(T)** | 33.05 |
| % deviance explained by full model = **19.2%** | Pop. Aged ≥ 65 | 18.38 |
| | Ag Employment | 7.10 |
| | Total Comp. Birds | 5.90 |
| N = 591 | SVI | 0.68 |
| **Southeastern** | **$R_0$(T)** | 59.41 |
| | **Total Comp. Birds** | 18.84 |
| % deviance explained by full model = **41.6%** | Edge Density | 10.93 |
| | SVI | 8.00 |
| | Ag Employment | 1.99 |
| N = 528 | Pop. Aged ≥ 65 | 0.82 |
| **Southwestern** | **$R_0$(T)** | 49.29 |
| | **Total Comp. Birds** | 16.53 |
| % deviance explained by full model = **23.8%** | Ag Employment | 14.17 |
| | SVI | 10.51 |
| | Pop. Aged ≥ 65 | 4.96 |
| N = 325 | Edge Density | 4.54 |
| **Western** | **$R_0$(T)** | 51.39 |
| | **Edge Density** | 23.12 |
| % deviance explained by full model = **45.0%** | Pop. Aged ≥ 65 | 11.10 |
| | SVI | 9.97 |
| | Total Comp. Birds | 3.18 |
| N = 661 | Ag Employment | 1.24 |

To support the selection of the final model, we include below a set of diagnostic visualizations: (i) covariates distributions and correlations (to assess multicollinearity), (ii) model diagnostics (to evaluate assumptions and fit), and (iii) partial dependence plots (to interpret the marginal effects of covariates on the outcome).

**(i) GAM covariate distributions.** Underlying data for county-level covariates.

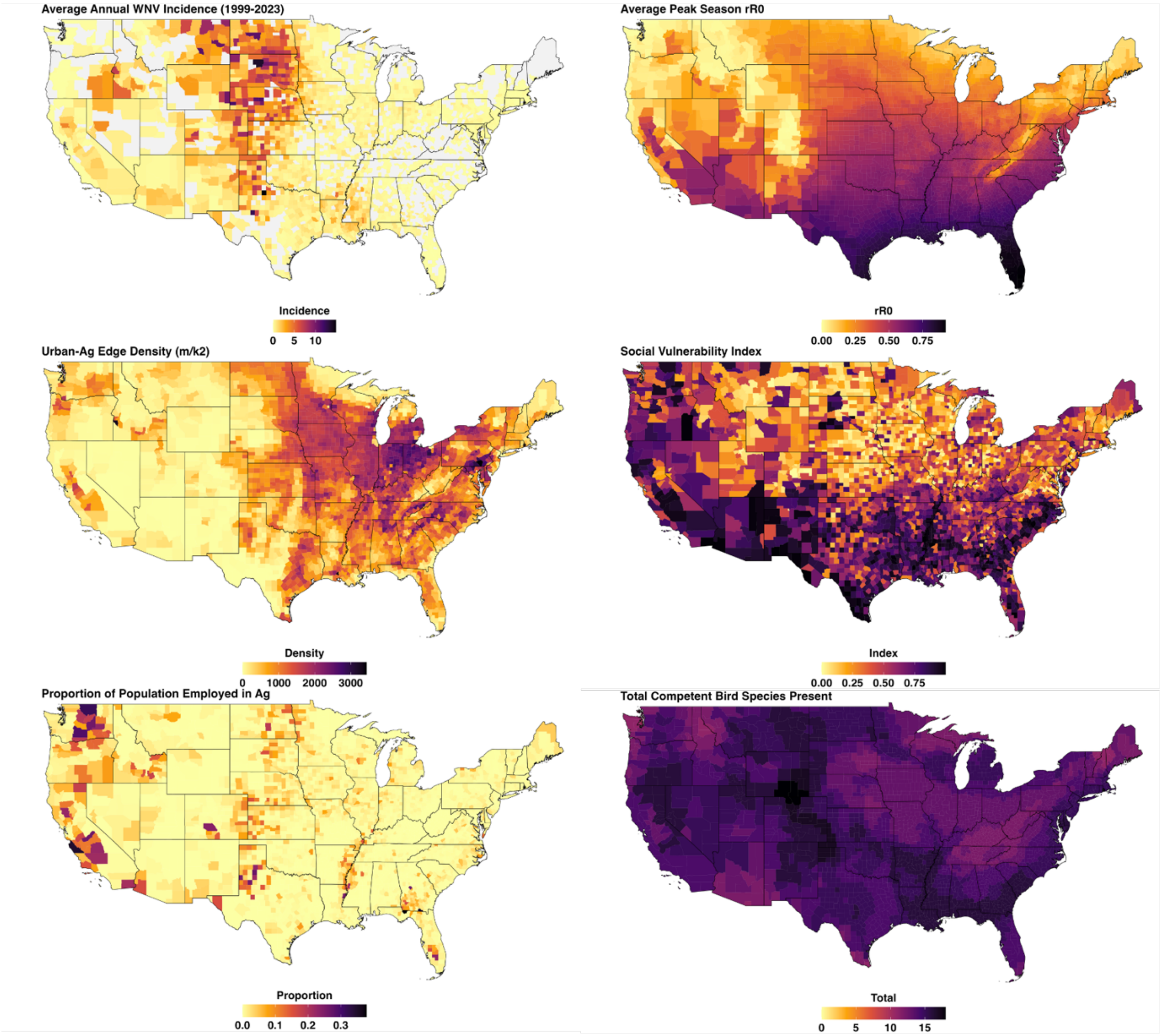


**(i) GAM covariate correlations.** Correlation values for covariates used in best fitted GAM model visualized with `ggcorrplot` package (21).

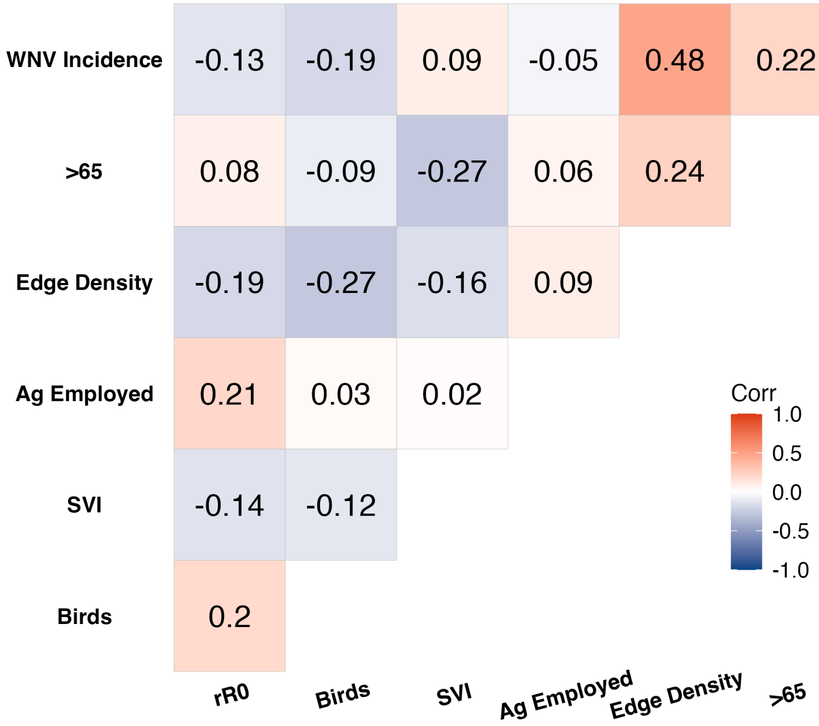


**(ii) GAM diagnostics.** Diagnostics for best fit GAM using the appraise() function from the \`gratia\` package (22).

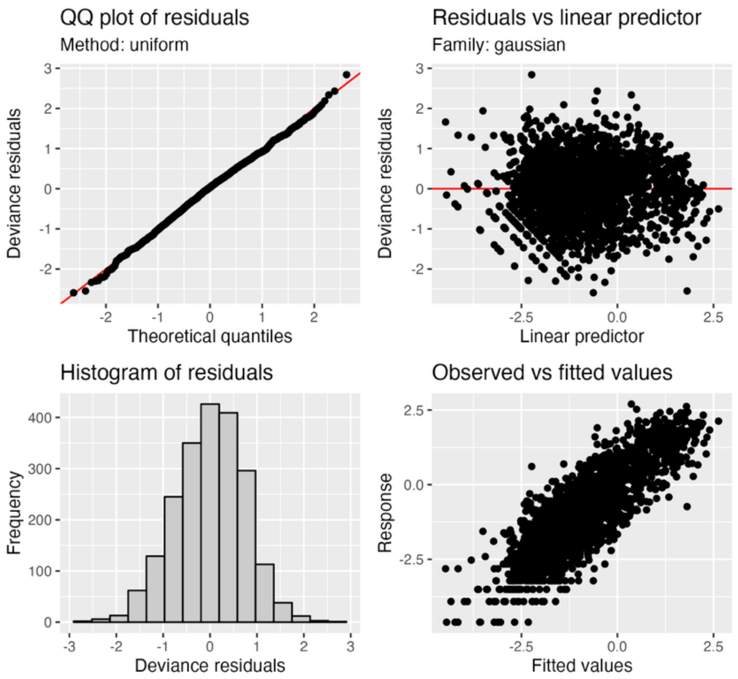


**(iii) GAM partial dependence plots.** Estimated effects for smooth terms on human WNVND incidence in the best fit GAM with 95% confidence intervals. Partial dependence plots were created with draw() function from the \`gratia\` package (22).

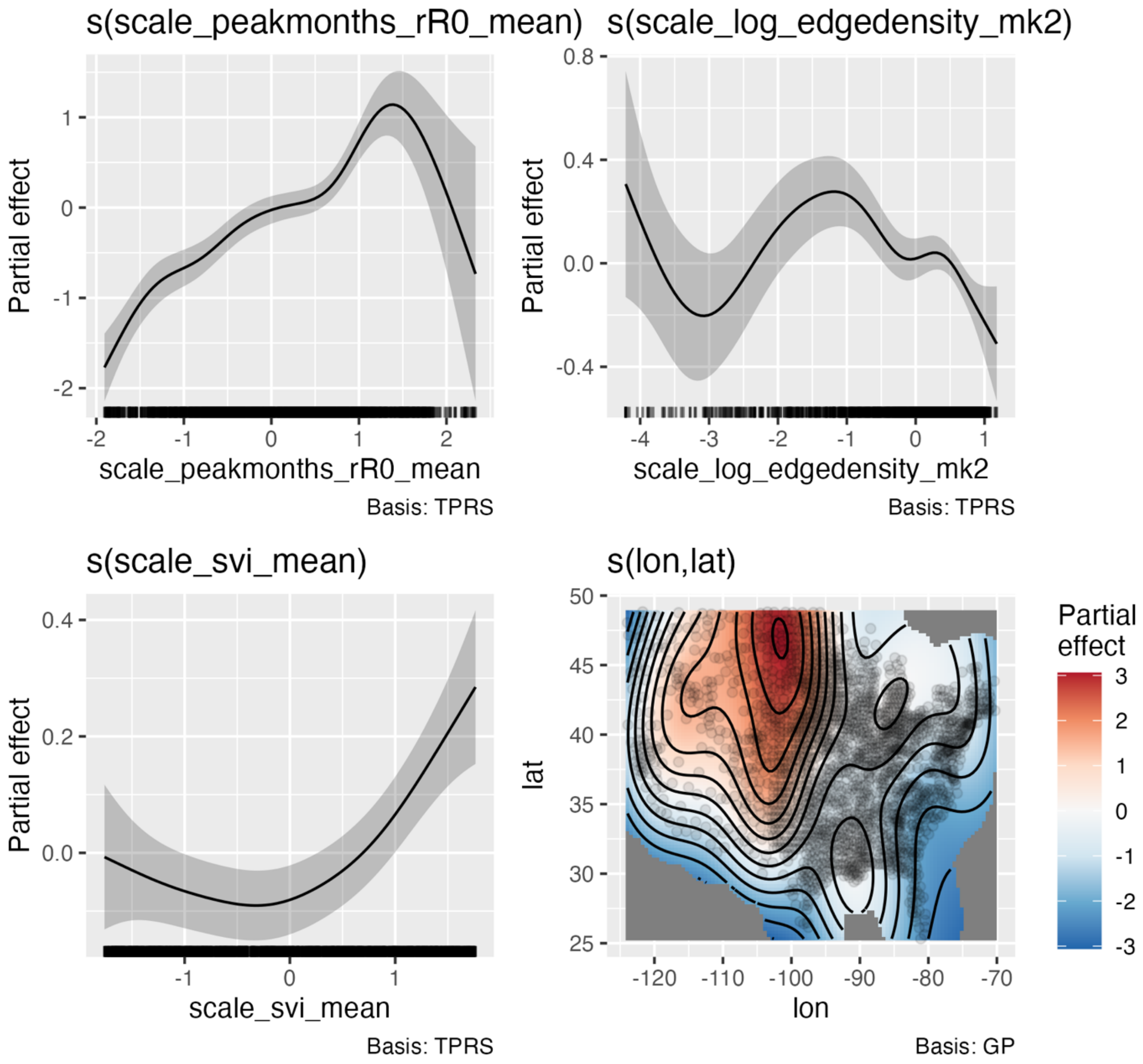
s(scale_peakmonths_rR0_mean)
Partial effect
scale_peakmonths_rR0_mean
Basis: TPRS
s(scale_log_edgedensity_mk2)
Partial effect
scale_log_edgedensity_mk2
Basis: TPRS
s(scale_svi_mean)
Partial effect
scale_svi_mean
Basis: TPRS
s(lon,lat)
lat
lon
Partial effect
Basis: GP

*S Text 2.2 Mechanistic temperature-dependent $R_0(T)$ analysis*

**Model specifications and species-specific fecundity adjustments**. The following $R_0(T)$ model equations and justifications are adapted from Shocket et al. (2020) (23). The model has been validated with independently collected human case data by Shocket et al. (2020) (23) and field-collected mosquito data by MacDonald et al. (2024) (24).

For *Cx. pipiens* and *Cx. tarsalis*, we use the following $R_0$ equation:

$$R_0(T) = \left( \frac{a(T)^2 bc(T) e^{\frac{-\mu(T)}{PDR(T)}} EFCG(T) a(T) EV(T) pLA(T) MDR(T)}{\mu(T)^3} \right)^{1/2}$$

For *Cx. quinquefasciatus*, we use the following $R_0$ equation:

$$R_0(T) = \left( \frac{a(T)^2 bc(T) e^{\frac{-\mu(T)}{PDR(T)}} ER(T) a(T) pO(T) EV(T) pLA(T) MDR(T)}{\mu(T)^3} \right)^{1/2}$$

*S Text 2.3 Projected habitat suitability*

**Projected land use analysis.** Initially, we aimed to incorporate forecasted land cover change, rather than static habitat suitability, into our temperature-dependent models to better identify potential future hotspots that account for both climate and land use change. Land cover projections were modeled for four scenarios consistent with the IPPC and downscaled using the forecasting scenarios of land use change (FORE-SCE) model (18). The land cover data were classified into 17 land-use land-cover categories, comparable to those used in the NLCD, and provided at a 250 m resolution. Projected land cover data for the years 2050 and 2100 were used to estimate habitat suitability for three mosquito species, *Cx. tarsalis*, *Cx. pipiens*, and *Cx. quinquefasciatus*. For each species, a habitat preference value between 0 and 1 was assigned to the 17 land-use land cover categories with higher values indicating a greater likelihood of suitability for that species. These values were informed by a recent species distribution map of each *Culex* species (19) and confirmed by an extensive review of the literature on mosquito habitat preferences and land cover associations (2,3,7,25–27). Using these species-specific preference values, categorical land use land cover categories were reclassified into continuous habitat suitability raster for each mosquito species. *Cx. tarsalis* was assigned a habitat suitability of 0.9 for agricultural land and 0.0 for developed land, while *Cx. pipiens* and *Cx. quinquefasciatus* were assigned 0.9 for developed land and 0.0 for agricultural land. Resulting map below of projected WNV risk metric based on projected temperature and land cover for mid-century (2045-2074) and late-century (2075-2100) and for each *Culex* species at a ~1.5 km spatial resolution.

However, for the final model we only use urban and agriculture land covers. We chose not to include forecasted land cover for all land cover types (codes 1-17) in the final analysis because of less certainty of how projected additional landcovers would be suitable for individual *Culex* species. For example, the projected suitability for *Cx. quinquefasciatus* suggested suitability in forested regions but failed to account for elevation constraints in the Rocky Mountain region (see maps below). Despite its limitations, we view this approach as a promising avenue for future work.

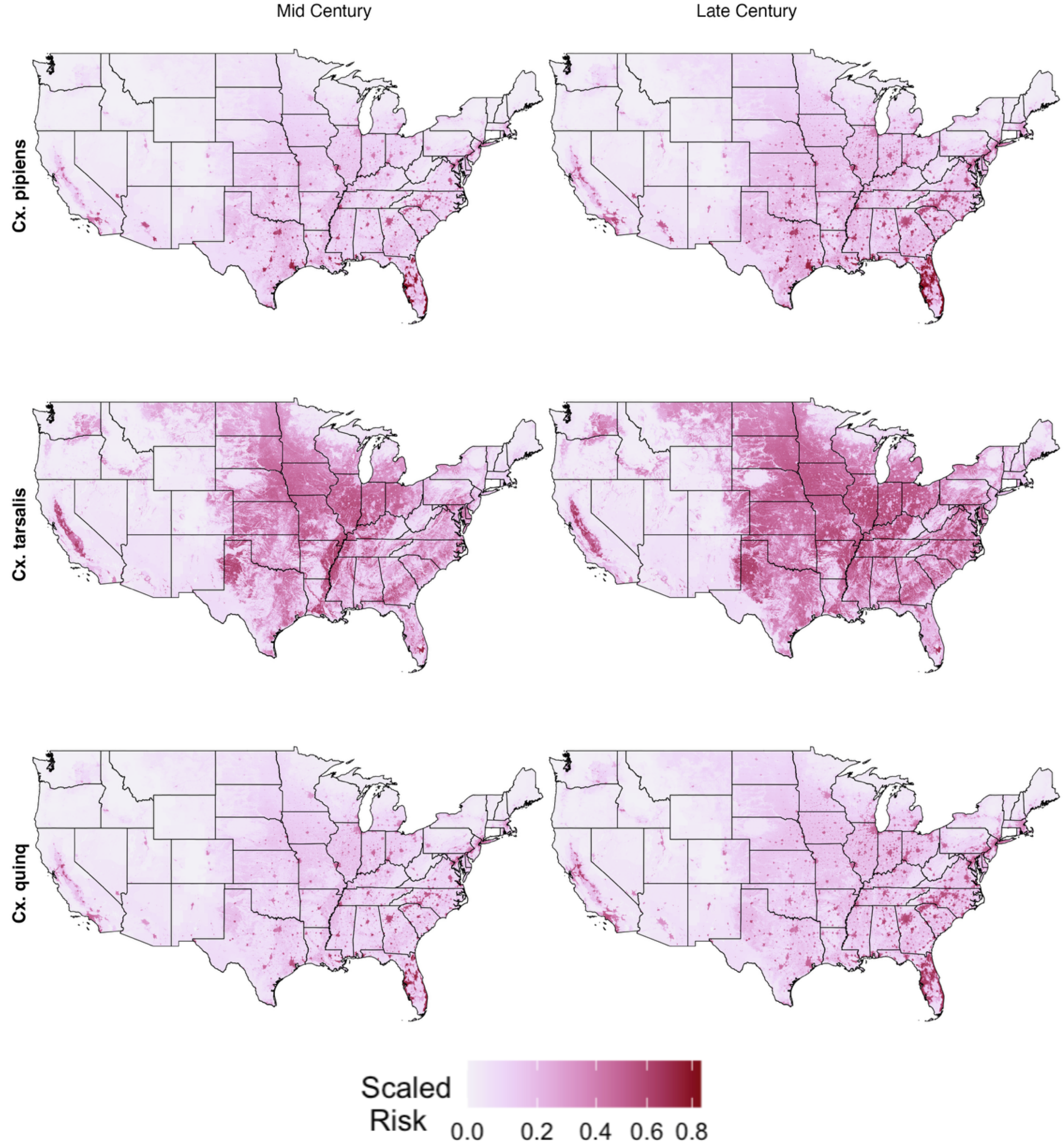


**Figure ST2.3**. Maps of projected scaled risk for mid-century (2045-2074) and late-century (2075-2100) under emission scenario SSP2-4.5 for each *Culex* species. Scaled risk is the product of temperature ($R_0(T)$) (scaled between 0-1) and land use suitability (0 or 0.9 depending on land cover type and species preference). Darker values represent areas with relatively high temperature and land use suitability. Land suitability in this figure is assigned for all land cover types (codes 1-17).

**Supplementary Text 3**. Estimating social vulnerability risk

*S Text 3.1 Evaluating the components of SVI*

To better understand the contribution of specific components of the Social Vulnerability Index (SVI), we selected six out of the 15 variables that comprise of the index that come from the American Community Survey (ACS) based on census-tract data from 2016-2020. Individual variables represent estimated total persons within a category. We hypothesized these variables to have plausible mechanistic links to increased WNV risk (Hypothesized mech.). These mechanisms include increased mosquito exposure, reduced access to healthcare, and barriers to prevention or mitigation efforts. Each variable was substituted individually into the full model in place of the full SVI and models were fit using the same GAMs. For each model, we report the model p-value, as well as the relative percent deviance explained by the individual predictor (i.e., the variable of interest), along with the range of deviance explained by all predictors within the fitted model calculated using the `gam.hp` package (20). We also include the total model deviance explained. This approach allows for clearer interpretation of each variable's explanatory power and alignment with hypothesized socio-ecological pathways.

| Model | Variable full name | Hypothesized mech. | p-value | % (range) |
|---|---|---|---|---|
| All themes | Social Vulnerability Index | Increase social vulnerability, increase risk to mosquito exposure and barriers to prevention efforts, as well as reduced access to healthcare | $5.67 \times 10^{-5}$ | Individual: 3.02 (2.21-75.61)<br>Model: 72.3% |
| POV150 | Persons below 150% poverty estimates | Lower housing quality may increase mosquito entry | $2.0 \times 10^{-16}$ | 20.32 (1.94-20.32)<br>Model: 74.2% |
| UNINSUR | Uninsured in the total civilian noninstitutionalized population estimate | Decreased access to care may increase risk of more severe disease | $2.0 \times 10^{-16}$ | 14.46 (2.02-66.96)<br>Model: 73.2% |
| LIMENG | Persons (age 5+) who speak English "less than well" estimate | Language barriers may reduce access to public health materials and response | 0.00093 | 5.27 (2.19 - 73.55)<br>Model: 72.1% |
| MUNIT | Housing in structures with 10 or more units estimate | Higher density housing may increase mosquito breeding habitat | $2.0 \times 10^{-16}$ | 9.33 (2.19 - 70.79)<br>Model: 72.8% |
| MOBILE | Mobile home estimate | May reflect lower housing quality may increase mosquito entry | $2.0 \times 10^{-16}$ | 19.12 (2.15-62.57)<br>Model: 73.6% |

| | | | | |
|---|---|---|---|---|
| CROWD | At household level (occupied housing units), more people than rooms estimate | Lower housing quality may increase mosquito entry | $2.0 \times 10^{-16}$ | 10.58 (2.13 - 69.8) Model: 72.8% |

*S Text 3.2 Additional covariates to measure social vulnerability*

In addition to the CDC's social vulnerability index (SVI), we examined more targeted indicators that directly relate to physical conditions that may affect residential exposure – such as the age of housing units (a proxy for the likelihood of damaged window screens) and the duration of power outages (as a proxy for periods without air conditions, during which residents may open windows or doors). For example, we used housing age and power outage duration in our GAMs, which were ultimately omitted due to their less interpretable results. For housing age, county-level estimates of the age of housing units were obtained from the US Census Bureau's American Community Survey (ACS) (Table ID: B25034, https://data.census.gov/table/ACSDT5Y2020.B25034?q=B25034). To estimate the proportion of older housing, the total number of housing units built in 1979 or earlier – including units categorized as built in pre-1939, 1940-1949, 1950-1959, 1960-1969, and 1970-1979 – was summed and divided by the total number of housing units in each county. These data are based on 2015-2020 ACS 5-year estimates. Housing unit age was used as a proxy for infrastructure vulnerability, under the assumption that older homes may be more likely to lack air conditioning (e.g., need to open windows/doors for cooling) or have deteriorated structural barriers (e.g., damaged window screens), potentially increasing indoor mosquito exposure. For power outage duration, county-level estimates of electricity outage per customer were derived from Brelsford et al. (2024)(28). We obtained data from https://figshare.com/articles/dataset/The_Environment_for_Analysis_of_Geo-Located_Energy_Information_s_Recorded_Electricity_Outages_2014-2022/24237376, specifically the estimates for 1) total number of utility customers per county as of 2022 (MCC.csv) and 2) total hours of electricity outage per customer per 2022 (eaglei_outages_2022.csv). We then calculated the average hours of electricity outage per customer per county. We used this calculation as a proxy for when traditional cooling methods (e.g., AC or fan) were not available to households so alternative means of cooling the household such as opening windows or doors would be necessary, and could potentially facilitate mosquito entry, thus creating residential risk. Although both metrics are proxies, we want to include metrics that may represent the physical processes driving WNV transmission, rather than broad social vulnerability indices.

*S Text 3.3 Validating SVI with other vulnerability metrics*

To assess whether the SVI correlates with other mechanistic variables not included in the composite, we examined its relationship with the percentage of homes built before 1980, a proxy for older or lower-quality housing that may facilitate indoor mosquito exposure. Across US counties, SVI showed a positive and statistically significant correlation with this housing-age metric (Spearman's rho = 0.19, $p < 2.2 \times 10^{-16}$), reinforcing the idea that higher social vulnerability may partially capture housing conditions conducive to indoor *Culex* exposure. We visualize a fitted relationship (method = "loess") between scaled SVI and % of houses built pre-1980, aggregated at county level.

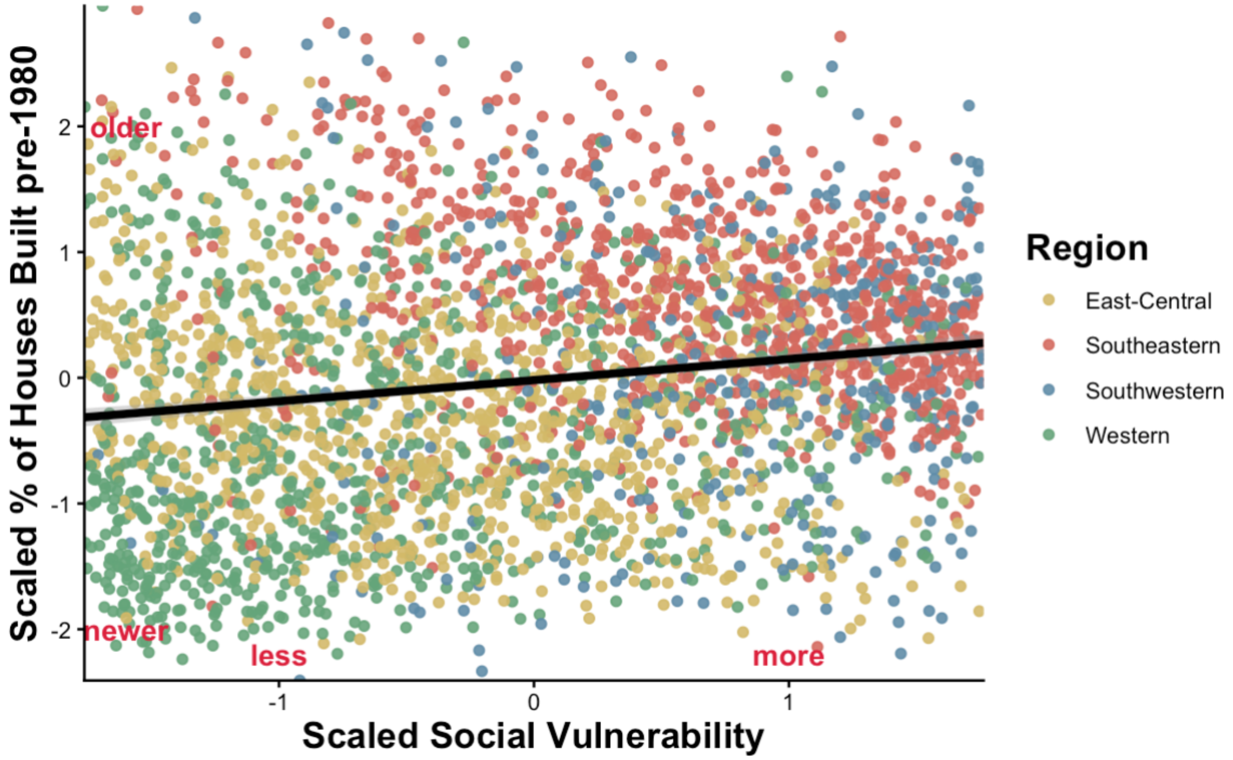

SUPPLEMENTARY TABLES

**Supplementary Table 1**. Regional classifications with contiguous United States based on Rochlin et al. (2019) Table 1 (2). Note that Washington, D.C. is included within a designated region. California (CA) is divided between two regions; see the footnote for details on county-level assignments.

| **Regions** | **States** |
|---|---|
| East-Central | CT, DC, DE, IL, IN, KY, MA, MD, ME, MI, NH, NJ, NY, OH, PA, RI, VA, VT, WI, WV |
| Southeastern | AL, AR, FL, GA, LA, MO, MS, NC, SC, TN |
| Western | CO, IA, ID, KS, MN, MT, ND, NE, NV, OR, SD, UT, WA, WY, CA[a] |
| Southwestern | AZ, NM, OK, TX, CA[b] |

[a]*CA county codes*: 001, 003, 005, 007, 009, 011, 013, 015, 017, 021, 023, 033, 035, 041, 043, 045, 047, 049, 051, 055, 057, 061, 063, 067, 075, 077, 081, 085, 087, 089, 091, 093, 095, 097, 099, 101, 103, 105, 109, 113, 115

[b]*CA county codes*: 019, 025, 027, 029, 031, 037, 039, 053, 059, 065, 069, 071, 073, 079, 083, 107, 111

**Supplementary Table 2**. Temperature-trait model parameters for the key *Culex* vectors of West Nile virus in the United States. Parameter values for the temperature-trait models were sourced from Shocket et al. (2020) (23).

***CX. PIPIENS***

| Trait | F(x) | $T_{min}$ | $T_{max}$ | q | Notes |
|---|---|---|---|---|---|
| Biting rate (a) | B | 9.4 | 39.6 | 0.000170 | |
| Vector competence (bc) | Q | 16.8 | 38.9 | 0.00305 | |
| Pathogen development rate (PDR) | B | 11.4 | 45.2 | 0.0000738 | |
| Eggs per female per gonotrophic cycle (EFGC) | Q | 5.3 | 38.9 | 0.598 | EFD = EFGC*a |
| Egg viability (EV) | Q | 3.2 | 42.6 | 0.00211 | |
| Prop. of larvae surviving to adulthood (pLA) | Q | 7.8 | 38.4 | 0.0036 | |
| Mosquito development rate (MDR) | B | 0.1 | 38.5 | 0.0000376 | |
| | **F(x)** | **m** | **z** | | |
| Lifespan (lf) | L | 4.86 | 169.8 | | |

***CX. TARSALIS***

| Trait | F(x) | $T_{min}$ | $T_{max}$ | q | Notes |
|---|---|---|---|---|---|
| Biting rate (a) | B | 2.3 | 32.0 | 0.000167 | |
| Transmission efficiency (b) | Q | 11.3 | 41.9 | 0.00294 | |
| Pathogen development rate (PDR) | B | 11.2 | 44.7 | 0.0000657 | |
| Eggs per female per gonotrophic cycle (EFGC) | Q | 5.3 | 38.9 | 0.598 | EFD = EFGC*a (used values for Cx. pipiens) |
| Egg viability (EV) | Q | 3.2 | 42.6 | 0.00211 | Used values for *Cx. pipiens* |
| Prop. of larvae surviving to adulthood (pLA) | Q | 5.9 | 43.1 | 0.00294 | |
| Mosquito development rate (MDR) | B | 4.3 | 39.9 | 0.0000412 | |
| | **F(x)** | **m** | **z** | | |
| Lifespan (lf) | L | 1.69 | 69.6 | | |

***CX. QUINQUEFASCIATUS***

| Trait | F(x) | $T_{min}$ | $T_{max}$ | q | Notes |
|---|---|---|---|---|---|
| Biting rate (a) | B | 3.1 | 39.3 | 0.0000728 | |
| Vector competence (bc) | Q | 4.2 | 45.2 | 0.00232 | Used values for *Cx. univittatus* |
| Pathogen development rate (PDR) | B | 19.0 | 44.1 | 0.0000712 | |
| Eggs per raft (ER) | Q | 5.0 | 37.7 | 0.0636 | EFD = ER*a |
| Prop. of females ovipositing (pO) | Q | 1.7 | 31.8 | 0.000667 | Switched from B to Q to allow for computation |
| Egg viability (EV) | B | 13.6 | 38.0 | 0.0047 | |
| Prop. of larvae surviving to adulthood (pLA) | Q | 8.9 | 37.7 | 0.00426 | |
| Mosquito development rate (MDR) | B | 0.1 | 38.6 | 0.0000414 | |
| | **F(x)** | **m** | **z** | | |
| Lifespan (lf) | L | 3.8 | 136.3 | Lifespan (lf) | L |

**Supplementary Table 3**. Additional information on data availability. This table summarizes metadata for all variables used in the analysis. “Variable ID” corresponds to the term used in the manuscript figures. “Variable details” provide additional information to locate each indicator and its associated data source. The “Temporal scale” column describes the reference period or time range covered by each dataset. Full source attributions are listed below the table.

| Source | Variable ID | Variable details | Temporal scale |
|---|---|---|---|
| 1 | WNV incidence | West Nile virus human neuroinvasive disease average annual incidence per 100,000 population at the county-level | 1999-2023 |
| 2 | Aged ≥ 65 | Percentage of the population aged 65 and older at the county-level | 2015-2020 |
| 3 | Land cover | 16-class land cover annual classifications at 30 m | 2020 |
| 4 | Bird community | Geographic bird species ranges and predicted habitats (individual polygons) | 2001 |
| 5 | Ag. workers | Proportion of the workforce employed in agriculture at the county-level | 2020-2023 |
| 6 | Social index | Social Vulnerability Index (SVI) which relates to demographic and socioeconomic characteristics at the county-level | 2018, 2020, 2022 |
| 7 | CMIP6 temp. | Temperature projections from the Coupled Model Intercomparison Project 6 (CMIP6), specifically the GFDL-CM4 global climate model, statistically downscaled to ~ 6 km using LOCAv2 (29,30) | 2015-2020 (current), 2045-2074 (mid-century), 2075-2100 (late-century) |
| 8 | *Culex* habitat | Predicted habitat suitability of predominant *Culex* mosquitoes across the Americas at 30 m (31) (netCDF files per species) | 1990-2020 |
| 9 | FORE-SCE | Projected land cover classifications from USGS Earth Resources Observation and Science Center using the forecasting scenarios of land-use change (FORE-SCE) model at 250 m (32) | 2020 (current), 2050 (mid-century), 2100 (late-century) |

[1]**ArboNET.** “West Nile virus human neuroinvasive disease average annual incidence per 100,000 population by county of residence, 1999-2024*” *Historic Data*, U.S. Centers for Disease Control and Prevention. https://www.cdc.gov/west-nile-virus/data-maps/historic-data.html. Accessed 8 August 2025.

[2]**US Census Bureau.** “65 years and over AGE AND SEX” *American Community Survey 5-year 2020*, US Census Bureau. https://data.census.gov/table/ACSST5Y2023.S0101?q=S0101&g=010XX00US$0500000. Accessed 8 August 2025.

[3]**National Land Cover Database (NLCD).** "Annual National Land Cover Database Collection" *National Land Cover Database*, US Geological Survey. https://www.mrlc.gov/data/land-cover-conus-6. Accessed 8 August 2025.

[4]**Gap Analysis Program (GAP)**. "Species Data" *Gap Analysis Program*, US Geological Survey. (https://gapanalysis.usgs.gov/apps/species-data-download/. Accessed 8 August 2025.

[5]**Bureau of Labor Statistics (BLS).** "Quarterly Census of Employment and Wages (QCEW)" *North American Industry Classification System (NAICS)*, Bureau of Labor Statistics. https://data.bls.gov/cew/apps/data_views/data_views.htm#tab=Tables. Accessed 8 August 2025.

[6]**Agency for Toxic Substances and Disease Registry (ATSDR).** "Social Vulnerability Index" *Place and Health – Geospatial Research Program*, US Centers for Disease Control and Prevention. https://www.atsdr.cdc.gov/place-health/php/svi/index.html. Accessed 8 August 2025.

[7]**Coupled Model Intercomparison Project 6 (CMIP6).** "LOCA2/NAmer/GFDL-CM4/0p0625deg/r1i1p1f1" *Historical, ssp245, ssp585*, LOCA2. https://cirrus.ucsd.edu/~pierce/LOCA2/NAmer/GFDL-CM4/0p0625deg/r1i1p1f1/. Accessed 8 August 2025.

[8]**Culex habitat.** "culexmaxentmodels" *Los Alamos National Laboratory* (*lanl)*, GitHub. https://github.com/lanl/culexmaxentmodels. Accessed 8 August 2025.

[9]**FORE-SCE.** "Conterminous United States Land Cover Projections – 1992 to 2100" *Forecasting Scenarios of Land-Use Change (FORE-SCE) under A2 scenarios*, US Geological Survey. https://www.usgs.gov/special-topics/land-use-land-cover-modeling/science/land-cover-modeling-methodology-fore-sce-model. Accessed 8 August 2025.
